# Biopsychosocial response to the COVID-19 lockdown in people with major depressive disorder and multiple sclerosis

**DOI:** 10.1101/2022.05.10.22274890

**Authors:** S Siddi, I Giné-Vázquez, R Bailon, F Matcham, F Lamers, S Kontaxis, E Laporta, E Garcia, B Arranz, G Dalla Costa, A.I Guerrero Pérez, A. Zabalza, M Buron, G Comi, L. Leocani, P Annas, M Hotopf, BWJH Penninx, M Magyari, P. S. Sørensen, X Montalban, G Lavelle, A Ivan, C Oetzmann, M K White, S Difrancesco, P Locatelli, DC Mohr, J Aguiló, V Narayan, A Folarin, R Dobson, J Dineley, D Leightley, N Cummins, S Vairavan, Y Ranjan, Z Rashid, A Rintala, G De Girolamo, A Preti, S Simblett, T Wykes, PAB members, I Myin-Germeys, JM Haro, the RADAR-CNS consortium

## Abstract

**Background:** Changes in lifestyle, finances and work status during COVID-19 lockdowns may have led to biopsychosocial changes in people with pre-existing vulnerabilities such as Major Depressive Disorders (MDD) and Multiple Sclerosis (MS).

**Methods:** Data were collected as a part of the RADAR-CNS (Remote Assessment of Disease and Relapse – Central Nervous System) programme. We analyzed the following data from long-term participants in a decentralized multinational study: symptoms of depression, heart rate (HR) during the day and night; social activity; sedentary state, steps and physical activity of varying intensity. Linear mixed-effects regression analyses with repeated measures were fitted to assess the changes among three time periods (pre, during and post-lockdown) across the groups, adjusting for depression severity before the pandemic and gender.

**Results:** Participants with MDD (N=255) and MS (N=214) were included in the analyses. Overall, depressive symptoms remained stable across the three periods in both groups. Lower mean HR and HR variation were observed between pre and during lockdown during the day for MDD and during the night for MS. HR variation during rest periods also decreased between pre-and post-lockdown in both clinical conditions. We observed a reduction of physical activity for MDD and MS upon the introduction of lockdowns. The group with MDD exhibited a net increase in social interaction via social network apps over the three periods.

**Conclusions:** Behavioral response to the lockdown measured by social activity, physical activity and HR may reflect changes in stress in people with MDD and MS.

## INTRODUCTION

In January 2020, the World Health Organization (WHO),(1) declared the new coronavirus SARS-CoV-2 epidemic, which caused COVID-19, to be a Public Health Emergency of International Concern. The first cases were identified in Wuhan, Hubei Province, China, in late December 2019. On March 11th 2020, the WHO declared the COVID-19 outbreak a pandemic (1).

When the virus appeared, several countries imposed a national lockdown to limit the spread of the virus, with restrictions varying from country to country. The government in Italy announced a nationwide lockdown on March 9th, 2020 (2).

The lockdown was implemented in Denmark and Spain one week later on March 13^th^ (3) and 14^th^ (4), respectively, but with differing restrictions. Other countries, such as the UK (5) and the Netherlands, implemented a lockdown in late March (6). The lockdown measures during the COVID-19 outbreak were expected to have adverse consequences, including financial losses, scarcity of basic supplies, family separations, and an increased perception of risk (7–10). This pandemic has increased the necessity to strengthen mental health systems; the cases of Major Depressive Disorders (MDD) increased globally by 27. 6% (11). Google Trends analysis revealed a massive rise in searches related to symptoms of depression and stress during the COVID-19 pandemic (12). These negative consequences could represent an even higher toll on people who already had a pre-existing mental illness (13). The psychological effects of “the COVID-19 pandemic reported were a reduction face to face social contacts (14), increased levels of loneliness (15–17), psychological distress (18), changes in depression levels (7), decreased self-esteem and sleep duration (14,19) and, in extreme cases, suicide (8).

The literature has widely documented the impact of stressful life events on the major depressive disorder (MDD) (20,21) and also on Multiple Sclerosis (MS) (22,23). MS disease has high comorbidity with depression (24). Consistent evidence shows high prevalence rates of depression (31%) and anxiety (22%) in MS (24). The restrictions required by the lockdowns may also have worsened depression symptoms in patients with MS. A review reported that individuals with MS are likely to feel depressed due to their psychosocial circumstances (25). They felt helpless, had low quality of social relationships, high levels of stress, and adopted maladaptive coping strategies. Psychosocial factors related to a lack of social support, adverse experiences, and life events may significantly impact women more than men (26). Although there is a large body of literature on the psychological effects of COVID-19, most of it has selected convenient samples and has used non-standardized instruments for the diagnosis (18). There is also limited information on the impact on individuals that suffer from depression or other chronic conditions; COVID-19 might have exacerbated pre-existing clinical symptoms in people with MDD and MS.

Remote measurement technology (RMT) allows to collect data in real-time and in a natural environment without high costs. It does not require face-to-face contact between the research team and participants. Decentralized (“virtual”) research with RMTs provides a vast quantity of data during over period without intruding into the daily life of research participants (27). The RADAR-CNS (Remote Assessment of Disease and Relapse – Central Nervous System) consortium developed the open-source mHealth platform RADAR-Base (28) to collect longitudinal data using RMT (phone and activity trackers), providing high-frequency data on depressed mood, self-esteem, speech, and cognition; and passively on heart rate, physical activity, sleep, and sociability. Previous analyses of this large sample were conducted to explore the impact of the pandemic on the behaviour (physical, sleep, social activity) focused on the differences by country (14) and another focusing exploring depression, self-esteem, and sleep in people with MDD (19), Both studies demonstrated the validity and utility of the RADAR-base to detect changes during the pandemic in the studied sample.

Considering the lockdown restrictions, increased levels of depression, reduced physical activity, and intensified social interaction via phone were expected. In this decentralized study, we also explored the photoplethysmography (PPG)-based Heart rate (HR) series provided by a commercial activity tracker.

The variability of HR is often used as an indicator of autonomic nervous system (ANS) regulation of the heart (29–31). HR can vary significantly over 24 hours in different conditions (32). In general, increased HR variability reflects a healthy ANS function that can respond to changing circumstances (33), while low HR variability is a sign of monotonously regular HR (34,35). HR variability is altered in people with depression (36) as they could have difficulties in psychological arousal regulation in the presence of emotional or environmental stressors (37). Dysfunctions in HR variability have also been found in people with MS (38,39). Alterations in HR could be an indicator of stress (40). Since MDD and MS have also been linked to alterations in HR, these alterations may have influenced their stress response.

The main objective of this study was to explore the impact of the lockdown, exploring the changes in depression level, physical activity, HR and sociality across three COVID-19 periods (pre-lockdown, during-lockdown and post-lockdown) and understand if changes depend on the depression severity before the pandemic in people with MDD and MS. We have also explored the impact of gender on the previous measures.

## METHOD

### Design

Digital data were collected as part of the international research consortium RADAR-CNS, a decentralized collaborative research initiative aiming to provide real-time multidimensional indicators of the clinical states of individuals with different health conditions: MDD, MS and epilepsy. This manuscript, we focused our analyses on two health conditions: MDD (41) and MS (42). A total of 1060 participants were recruited in 5 European countries: people with Major Depressive Disorder (Netherlands, Spain and the UK) and people with Multiple Sclerosis (Italy, Denmark and Spain).

The recruitment and follow-up started in November 2017 and finished in March 2021. For this study, we focused on the following periods:

- During lockdown: we chose the entire period of the national lockdown in each country.
- For the pre-lockdown phase, we chose the period immediately before the first restrictive measure with the same duration as the total national lockdown. We chose the period immediately posterior to the national lockdown for the post-lockdown phase with the same duration.

The following dates were considered to define the period:

- Denmark: lockdown began 13/03/2020, and post-lockdown started 16/04/2020
- Italy: lockdown began 09/03/2020, and post-lockdown started 19/05/2020
- Spain: lockdown began 14/03/2020, and post-lockdown started 05/05/2020
- The Netherland: 15/03/2020 post-lockdown and started 12/05/2020
- UK: lockdown began 23/03/2020, and post-lockdown started 12/05/2020

Different restrictions were implemented across countries. Italy and Spain had the more strict restrictions on gatherings and school closures (only geographically targeted). The requirement of staying at home except for essential trips and the cancellation of public events was implemented in all countries except Denmark, where the measure was only recommended. Places of work were required to close in some sectors in Spain, the United Kingdom, and Denmark, and were required to close for all but public services (stores, libraries, museum etc.) in the Netherlands and Italy. Public transport was recommended to close in Italy, Spain, and Denmark.

### Procedure

All local Ethics Committees approved the protocol of the institutions involved in the study, and participants provided written consent before enrollment (See (41) for MDD and (42) for MS).

### Instruments

We analyzed the following features derived from the open-source mHealth platform RADAR-Base (28). RADAR-base is developed as a modular application that includes active and passive apps for data collection. The active app includes questionnaires on mood, self-esteem, cognitive function and speech tasks. While the passive app that did not require active engagement, continuously collects data on a 24/7 basis through a smartphone and a Fitbit device, which includes phone usage, location, Bluetooth, heart rate and physical activity. For Fitbit devices (Fitbit Inc, San Francisco, CA, USA), Fitbit Charge 2/3 devices were given to participants. Participants were asked to wear this wearable device on their wrist of the non-dominant hand for the duration of the follow-up across cohorts, which provided ongoing information about physical activity and HR derived from the accelerometer and the PPG signals, respectively. This device was selected by the RADAR-CNS project members, involving the patients, who decided together to use a commercially available device which is minimally intrusive and easier to use (43–46). For this study we focused on the analyses of the heart rate, physical activity captured using a wearable device, and social activity and depression symptoms were captured by using a Smartphone (Table 1).

**Table 1.** Variables assessed in the study.

|  |  |  |
| --- | --- | --- |
| **Clinical and social activity variables derived from the Apps** |  |  |
| **Depression Level** | PHQ-8 score | PHQ-8 score last observation before pre-lockdown phase. |
|  |  | Score range = 0-24 |
| | | Cut-off score $\geq 10$ moderate to severe depression |
| **Social activity** |  |  |
| *Contacts* | Daily mean of contacts | number of all contacts per day |
| *Social interactions* | Sum of interaction and short cut with the social apps | Number of interactions with social apps (whatsapp, facebook, twitter and others) used to make calls, read and send messages in one day |
| **Physiological features (derived from Fitbit device)** |  |  |
| **Physical activity** |  |  |
| **Heart rate parameters** |  |  |
| *Mean HR daily* | Mean of HR during the day | Mean HR across all day |
| *HR variation /day* | Std of HR during the day | Standard deviation of HR across all day. |
| *Resting mean HR day* | Mean of HR during the resting period across all day | Resting of mean HR across all day |
| *Resting HR variation / day* | Std of HR during the resting period across all-day | Std of HR during sedentary periods, identified by activity level = sedentary periods and number of steps = 0 |
| *Resting mean HR at night* | Mean of HR at rest during the night | Mean of HR during sedentary periods, identified by activity level = sedentary and number of steps = 0 only during night time (0:00-05:59) |
| *Resting HR variation at night* | Std of HR variation at night | Std of HR |
| *Steps* | Mean of steps | Mean of steps per minute along all day. |
| *Sedentary state* | “Sedentary” Minutes | The number of minutes in a day for which the physical activity was classified as “sedentary”. |
| *Light, Moderate and Vigorous activity* | “Active” Minutes | The number of minutes in a day for which the physical activity was classified as “lightly” or “moderate” and “vigorous activity” |

### Depressive symptoms

Depressive symptoms were assessed with the Patient Health Questionnaire (PHQ-8), representing a valid instrument to identify depression severity in MDD and MS (47–49). Participants with MDD were asked to complete the PHQ-8 (47) every two weeks and participants with MS, every six weeks through an app (active RMT)(28) installed on an Android smartphone. The responses for each item vary from 0= “not at all to 3 =“nearly every day”. The PHQ-8 score ranges from 0 to 24 (increasing severity). A cut-off score of ≥10 is the most recommended cut-off point for “clinically significant” depressive symptoms, which means that the participant is likely to meet diagnostic criteria for a depressive episode (or moderate and severe depression) in the previous two weeks. Ratings below 10 are usually defined as an asymptomatic state or sub-threshold (no or mild depression). Internal consistency was calculated with Cronbach’s alpha, and it was =0.91, for MDD and 0.88 for MS during the different assessments.

### Social activity

Participants installed another app (passive RMT app) that collected data on behavioural activity via smartphone sensors including social activity such as the numbers of contacts and app interaction. The data were collected continuously. *Social contacts* represent the number of new contacts added daily to the list of contacts concerning the previous measurement. The *social interaction* is represented by the sum of the number of interactions with social apps used to make calls, read and send messages per day through apps (i.e., Facebook, Messenger, Reddit, Skype, Telegram, Twitter, Viber, WhatsApp, etc.) measuring the activity of the user in these platforms.

### Heart rate

In this study, we used the heart rate (HR) parameters that were reported to be related to depression severity in a previous study (Siddi et al. submitted). Briefly, for assessing the HR profile, the mean level and standard deviation of HR were computed for the whole day, during resting/sedentary periods during the day and during resting/sedentary periods at night. This device was previously proven to measure HR accurately (31,50–52).

### Physical activity and sedentary levels

Sedentary levels are estimated based on the mean number of steps per minute during the day and physical activity is classified as sedentary, light, moderate and vigorous by proprietary algorithms of the Fitbit device. Physical activity was calculated by taking into account the time during which the individual was wearing the Fitbit. Periods without HR values are excluded from wear-time duration. Daily mean number of steps is a measure of global activity. Sedentary, light, moderate and vigorous activity were selected due to their strong association with depression (53–55) and during the pandemic (i.e. (56,57).

#### Statistical analysis

First, we described the sociodemographic characteristics reporting frequencies and percentages for categorical variables, mean and standard deviation or medians with interquartile ranges (IQR) for continuous variables, as appropriate. We computed the average of each of the daily parameters (HR, physical and social activity) in the week before the PHQ-8 assessment across each pandemic-related period.

Linear mixed-effects regression analyses for repeated measures (depressive symptoms, HR, physical activity and social activity) were conducted to assess the changes in the mean total score of each daily parameter among the three periods: pre-, during and post-lockdown (defined for each country).

We then added the baseline depression severity into each model (Depression severe or moderate=1, No depression or mild=0, using a PHQ-8 cut-off ≥10). The baseline depression severity was calculated from the last PHQ-8 measurement before the pre-lockdown period, so it is generally in the range of the previous two weeks prior to that period. We included a three-way interaction period for baseline depression severity and gender, as exposure variables, to investigate whether the rate of change in the outcome variables over time varied according to depression severity and gender. All models incorporated a random effect for participants. All the analyses were conducted with R, packages Nonlinear Mixed Effects Models (nlme) (58) and Estimated Marginal Means (emmeans) (59).

## RESULTS

A total of 469 individuals, of which 71% were women, were included in the analyses, 255 people with MDD and 214 with MS (Table 2). We excluded participants with missing observations of PHQ-8 in the pre-lockdown interval (people with no data in the pre-lockdown as they were enrolled later or missed the assessment). Despite the large amount of digital data collected thanks to the RMT that monitors participants per day over a long period, missing values may occur due to technical problems and daily life (levels of completeness have been reported in the supplementary materials Table S1.). For more details on available data in the MDD cohort (60). The mean age in the sample was 46.6 years old (SD=13.3). Half of the individuals in the MDD had no partner (50.6%), while in the group with MS, most had a partner (65.6%). Years of education were higher in the MS group compared with the MDD group.

**Table 2.** Sample and feature characteristics.

|  | **MDD (N=255)** | **MS (N= 214)** |
| --- | --- | --- |
| **Age (Mean, SD)** | 47.73 (15.42) | 44.77 (9.94) |
| **Gender (Women, N %)** | 190 (74.51) | 144 (67.29) |
| **Marital status** |  |  |
| **Without partner (Single/ Separated Divorced/Widowed)** | 129 (50.59%) | 73 (34.43%) |
| **(Partner/Married** | 126 (49.41%) | 139 (65.57%) |
| **Education year mean (SD)** | 15.01 (6.15) | 16.91 (5.91) |
|  | *Median (IQR)* | *Median (IQR)* |
| **PHQ-8** | 9 (10) | 5 (6) |
| **Physical activity** |  |  |
| Steps | 4.22 (5.03) | 4.53 (4.8) |
| Sedentary | 949.36 (385.89) | 925.29 (322.57) |
| Light activity | 138.07 (121.89) | 166.29 (150.07) |
| Moderate activity | 8.36 (17.14) | 8.43 (18.14) |
| Vigorous activity | 6.71 (20.86) | 3.86 (11.71) |
| **HR parameters** |  |  |
| mean HR day | 74.87 (11.22) | 76.72 (9.86) |
| std HR day | 12.46 (3.95) | 12.87 (4.11) |
| Resting mean HR day | 72.82 (11.03) | 75.44 (9.79) |
| Resting stdHR day | 11.14 (4.18) | 12.42 (3.79) |
| Resting mean HR at night | 66.53 (11.66) | 67.04 (10.58) |
| Resting stdHR at night | 4.93 (2.15) | 5.43 (3.03) |
| **Social activity parameters** |  |  |
| Social contacts | 136 (163) | 258 (344) |
| Social interactions | 5 (10) | 6 (13) |

**Note:** Note: **PHQ-8**= depression severity **Steps**= mean of steps per day, **Sedentary** = The number of minutes per day classified as a sedentary. **Light, moderate and vigourous activity**: the number of minutes per day classified as “lightly” or “moderate” and “vigorous “activity”, , **mHR day**=mean HR during 24h, **std HR** = standard deviation of HR during 24h, **restingHR day**= HR at rest during 24h, **Resting HR at night**= HR at rest during the night (0:00-05:59), **social contacts** = number of contacts in the agenda, **social interactions** = interaction trough the social apps

### Summary of findings in MDD

Table 3 displays the main findings across the three periods (pre, during and post-lockdown) in individuals with MDD. Overall, mean depressive symptoms remained stable across these periods.

**Table 3:** Estimated mean differences in each outcome between periods in MDD (Linear mixed effects model)

|  | pre- and during- lockdown<br>(95% CI, *p*-value) | pre and post- lockdown<br>95% CI, *p*-value) | during- and post-lockdown<br>95% CI, *p*-value) |
| --- | --- | --- | --- |
| **PHQ-8** | 0.24 (-0.32 to 0.81) | 0.03 (-0.58 to 0.64) | -0.22 (-0.78 to 0.35) |
| **Steps** | **1.32 (0.91 to 1.73) \*\*\*** | **0.79 (0.33 to 1.26)\*\*** | **-0.52 (-0.99 to -0.061) \*** |
| **Sedentary** | **52.0 (0.59 to 103.4)\*** | **73.7 (16.59 to 130.8)\*\*** | 21.7(-28.040 to 71.5) |
| **Light activity** | **38.06 ( 25.9 to 50.21) \*\*\*** | **30.35 (17.1 to 43.55)\*\*\*** | -7.71(-19.4 to 3.99) |
| **Moderate activity** | **4.24 (1.97 to 6.51) \*\*\*** | **2.94 (0.32 to 5.56)\*** | -1.30 (-3.02 to 0.42) |
| **Vigorous activity** | **3.45 (1.29 to 5.61) \*\*\*** | 1.31 ( -0.93 to 3.55) | **-2.14 (-4.28 to -0.004) \*** |
| **Mean HR day** | **1.38 (0.78 to 1.99)\*\*\*** | 0.43 (-0.29 to 1.14) | **-0.95 (-1.69 to -0.22)\*\*** |
| **std HR day** | **0.75 (0.37 to 1.14)\*\*\*** | **0.44 (0.039 to 0.84)\*** | -0.32 (-0.77 to 0.13) |
| **Resting meanHR day** | -0.037 (-0.64 to 0.57) | **-1.25 (-1.98 to -0.51)\*\*\*** | **-1.21 (-1.94 to -0.48)\*\*\*** |
| **Resting stdHR day** | **-0.72 (-1.24 to -0.19)\*\*** | **1.45 (-2.00 to 0.93) \*\*\*** | **-0.75 (-1.23 to 0.27) \*\*\*** |
| **Resting meanHR at night** | 0.31 (-0.40 to 1.03) | 0.53 (-0.23 to 1.29) | 0.21 (-0.39 to -0.82) |
| **Resting stdHR at night** | 0.065 (-0.21 to 0.34) | -0.06 (-0.38 to 0.26) | -0.12 (-0.44 to 0.19) |
| **Social contacts** | **-7.30 (-12.6 to -2.04)\*\*** | **-11.85 (-19.31 to -4.36)\*\*** | **-4.55 (-65.8 to -56.72)\*** |
| **Social interactions** | **-2.42 (-3.64 to -1.20)\*\*\*** | **-1.05 (-2.11 to 0.01)\*** | **1.37 (0.138 to 2.60)\*** |

**Note:** PHQ-8= depression severity, **mHR day**=mean HR during 24h, **std HR** = standard deviation of HR during 24h, **resting mHR day**= mean HR at rest during 24h, **Activity mHR day** = mean HR during activity period during 24h, **Resting mHR at night**= mean HR at rest during the night (0:00-05:59), **Steps**= mean of steps per day, **Sedentary** = The number of minutes per day classified as a sedentary. **Light, moderate and vigorous activity**: the number of minutes per day classified as “lightly” or “moderate” and “vigorous activity”**social contacts** = number of contacts in the agenda, **social interactions** = interaction trough the social apps
\* $p < 0.05$ ; \*\* $p < 0.01$ ; \*\*\* $p < 0.001$

MDD group reduced their physical activity during the lockdown as measured by the number of steps (p<0.001), and it increased post-lockdown (p<0.001) again. A similar pattern was found for light, a moderate and vigorous activity that reduced between the pre-lockdown and the lockdown (p<0.001). Light and moderate activity also decreased between the pre-and post-lockdown period (p<0.05). All individuals reduced their sedentary levels from pre-lockdown to post-lockdown (p<0.01).

During the day, mean HR and HR variation reduced between pre and during lockdown (p<0.001) while resting HR variation increased. Mean HR during all day (p<0.001) and resting state (p<0.01) increased again between lockdown and post-lockdown. No differences were found for resting HR at night.

We found a reduction of the HR variation (stdHR) all day (p<0.05), especially during the resting state (p<0.001) and an increase in mean HR at resting period (resting mHR) (p<0.001) during the day.

Overall, the social activity varied significantly during these periods. As we expected, the overall group intensified the social interactions through the apps, especially between pre and during the lockdown (Table 3).

### Differences in each outcome between no or mild depression vs moderate or severe depression at each period and interaction with gender

Figure 1 reports the differences between the groups with different depression severity (moderate or severe depression versus N=124 no or mild depression at baseline N=129).

**Figure 1.**
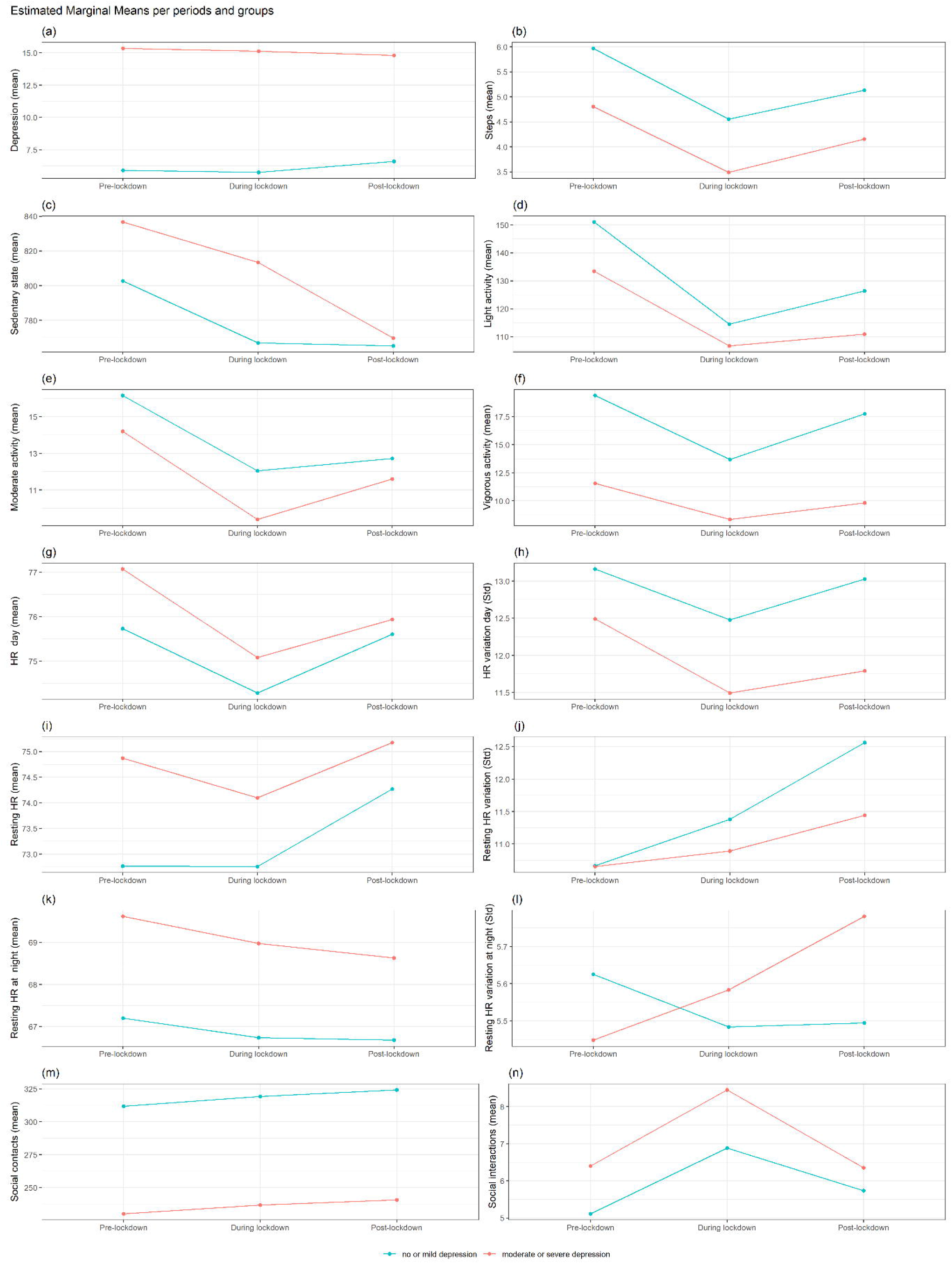
Interactions trajectories by depression status in MDD group

The group with moderate-severe depression at baseline reported higher depression levels in the three periods (p <0.0001) (Supplementary Materials Table S2)(Figure 1a). In the group with severe depression at baseline, women decreased the level of depression between pre and during lockdown compared to men (p <0.05).

No differences emerged for HR by depression severity pre and during the lockdown. The group with moderate or severe depression before the pandemic, compared to no or mild depression, reported lower HR variation all day (p<0.05) and at the resting state (p< 0.05) during the post-lockdown. Regarding the resting HR at night, in the group with higher depression before the pandemic, men decreased resting mean HR during the night between pre and post-lockdown compared to women (p<0.05).

The group with moderate or severe depression, compared to the mild or no depression group, reported a lower mean of steps (Figure 1b) during pre and lockdown and vigorous activity during all periods (Figure 1f). No significant differences emerged for social activity (Figure 1m and n) (p>0.05).

### Summary of findings for MS

Table 4 displays the main findings across the three periods (pre, during and post-lockdown) in the group with MS. In general, mean depressive symptoms remained stable for the MDD group across these periods.

**Table 4:** Estimated mean differences in each outcome between periods in MS.

|  | **pre- and during-lockdown (95% CI, *p*-value)** | **pre and post-lockdown 95% CI, *p*-value)** | **during- and post lockdown 95% CI, *p*-value)** |
| --- | --- | --- | --- |
| **PHQ-8** | -0.07 (-0.62 to 0.47) | -0.191 (-0.79 to 0.41) | -0.117 (-0.66 to 0.43) |
| **Steps** | **1.36 (0.82 to 1.89) \*\*\*** | 0.44 (-0.04 to 0.93) | **-0.91 (-1.35 to -0.48) \*\*\*** |
| **Sedentary** | -7.33 (-74.3 to 59.7) | -4.21 (-76.0 to 67.6) | 3.12 (-63.4 to 69.6) |
| **Light activity** | **35.5 (17.0 to 53.9) \*\*\*** | **12.7 (-9.0 to 34.49)** | **-22.7 (-41.8 to -3.67) \*** |
| **Moderate activity** | **3.72 (0.069 to 7.36) \*** | 2.71 (-0.90 to 6.33) | -1.01 (-4.30 to 2.30) |
| **Vigorous activity** | 2.17 (-1.11 to 5.46) | 1.14 (-2.22 to 4.49) | -1.04 (-5.01 to 2.93) |
| **mHR day** | **2.14 (1.26 to 3.02) \*\*** | **1.38 (0.44 to 2.32) \*\*** | -0.76 (-1.57 to 0.05) |
| **std HR day** | **1.10 (0.56 to 1.63) \*\*\*** | **0.80 (0.32 to 1.28)\*\*** | -0.30 (-0.85 to 0.25) |
| **Resting mHR day** | **1.67 (0.78 to 2.57) \*\*\*** | 0.64 (-0.28 to 1.57) | **-1.03 (-1.85 to -0.21) \*\*** |
| **Resting stdHR day** | **0.98 (0.48 to 1.49) \*\*\*** | **0.46 (0.014 to 0.91) \*** | -0.53 (-1.07 to 0.020) |
| **Resting mHR at night** | **1.20 (0.02 to 2.39) \*** | **1.37 ( 0.16 to 2.58) \*\*** | 0.16 (-1.04 to 1.37) |
| **Resting stdHR at night** | **0.57 (0.02 to 1.15) \*** | 0.12 (-0.29 to 0.53) | -0.44 (-1.01 to 0.11) |
| **Social contacts** | **-9.17 (-15.7 to -2.61) \*\*** | -18.01 (-44.4 to 8.40) | -8.84 (-37.2 to 19.55) |
| **Social interactions** | -1.92 (-4.15 to 0.31) | -1.01 (-3.46 to 1.44) | 0.91 (-2.33 to 4.14) |

**Note:** PHQ-8= depression severity, **mHR day**=mean HR during 24h, **std HR** = standard deviation of HR during 24h, **resting mHR day**= mean HR at rest during 24h, **Activity mHR day** = mean HR during activity period during 24h, **Resting mHR at night**= mean HR at rest during the night (0:00-05:59), **Steps**= mean of steps per day, **Sedentary** = The number of minutes per day classified as a sedentary. **Light, moderate and vigorous activity**: the number of minutes per day classified as “lightly” or “moderate” and “vigorous “activity”**social contacts** = number of contacts in the agenda, **social interactions** = interaction through the social apps
\* p<0.05; \*\* p<0.01; \*\*\* p < 0.001

As measured by the number of steps, activity was reduced during the lockdown and increased from the lockdown to post-lockdown (p<0.001). This was also observed for light activity between the pre-lockdown and during the lockdown (p<0.001) and increased again between the lockdown and the post-lockdown (p<0.05). Regarding HR, the most evident changes were observed between pre-lockdown and during the lockdown, and all HR parameters, either all day (p<0.001) or at night (p<0.05) were reduced.

The resting mean HR increased after the lifting of lockdown, as evidenced when comparing during- and post-lockdown periods (p<0.01). We found a reduction of the HR variation (stdHR) all day (p<0.05), especially during resting periods (p<0.001), and reduced mean HR during the night (p<0.01)

In the whole group of MS, the social activity remained stable, except for the social contacts, which increased from pre-lockdown to lockdown (p<0.001) (Table 4)

#### Differences in each outcome between no or mild depression vs moderate or severe depression at each period and interaction with gender

Figure 2 and Table S3 (Please see supplementary materials) reported the differences between the groups with different depression severity before the pandemic (moderate or severe depression versus N=53 no or mild depression at baseline N=152) across the three periods.

**Figure 2.**
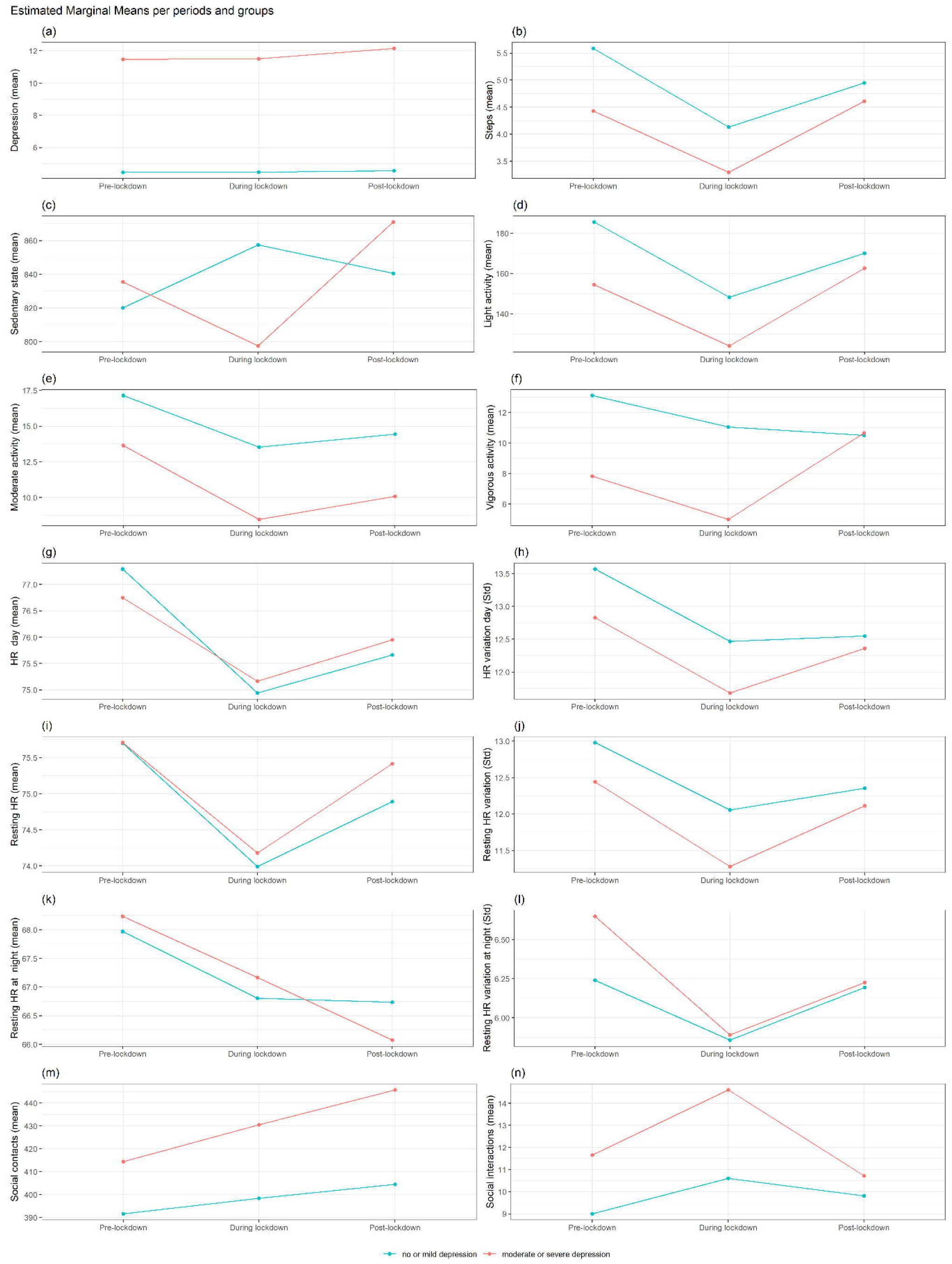
Interactions trajectories by depression status in MS group

No significant changes emerged across all periods by depression severity, and no impact of gender in the studied variables. The group with moderate or severe depression reported higher depression levels in all three periods (p<0.001) (Figure 2a).

The group with lower depression severity before the pandemic reported a higher mean of steps during the pre-lockdown (p<0.05) (figure 2b) compared to the other group. No differences between the groups for light, moderate and vigorous activity (figure 2c-f) and for social activity were found. No significant differences in gender emerged across these periods.

## DISCUSSION

Millions of people in different countries were confined in their homes due to the first wave of the COVID-19 pandemic, which has led to unprecedented social, personal and psychological experiences. Various factors, such as the closure of schools, interruption of work in some cases, closure of recreational gyms, the restrictions on seeing relatives, and the recommendation or obligation to remain at home, drastically changed regular routines and may have potentially increased levels of depression. Long term exposure to stress in individuals with long-term physical and mental health conditions can negatively affect mental health and emotional well-being (61). The majority of the publications in the literature explored the psychosocial effect of the lockdown on the general population (16,62). The RADAR-CNS project offered the opportunity to explore the biopsychological and social impact of the lockdown and pandemic situations on the participants with MDD and MS who participated before and after the lockdown. The present work provides essential findings on psychosocial response measured by depression severity, social activity, physical activity and HR as a proxy measure of stress due to the lockdown in people with MDD and MS.

### Findings on participants with MDD

Overall, the depressive symptoms remained stable across these periods. Previous studies have observed that lockdown did not cause changes to depression levels in people who were already depressed before pandemic (19,63,64). However, depression in women with more severe depression decreased from pre-lockdown to lockdown compared to men, though this did not occur after the lockdown. The closure of education centres and nursery schools led to an increase in children’s care and the volume of household chores. This situation may have contributed to more significant differences between women and men during the lockdown in different communities (18,65–69).

Although women may experience higher stress levels due to their greater responsibility, the ability to use coping strategies could be related to a better adaptation to new stressful situations than men (70). Another explanation might be that increasing social support during the pandemic may have enhanced women’s resilience (62). Lockdown measures have contributed to increased social isolation and feelings of loneliness (15–17,71). This could be one of the reasons why people with MDD showed intensified social activity measured through the social interactions with the apps and the number of contacts across the three periods, especially during the lockdown. This aligns with our previous work (14), which demonstrated an increased proxy for social activity measured by Bluetooth connections in during the lockdown. The group with MDD showed a reduced mean HR and HR variation during lockdown to the pre-lockdown period. A previous study showed that HR reduced during the lockdown (72), probably due to lower activity. We observed an increased resting mean HR between the lockdown and post-lockdown. From the pre- to post-lockdown, the patients with MDD had decreased the HR variation and increased the mean HR during the rest. Significant HR variation reflects the capacity to adapt to new situations; depression was found to be associated with low HR variability in previous studies, especially during the resting period (36,73,74). At the same time, we did not observe changes in resting mean HR and HR variation during the night. In the group with more severe depression severity before the pandemic, men’s resting mean HR decreased during the night from pre to post-lockdown compared to women. In general, men have a lower resting mean HR compared to women (75), though these differences may also vary with sleep quality, age and other factors such as the body mass index (76).

As expected, all groups reduced their physical activity (all intensities) during the lockdown. Light and moderate activity also decreased from pre to post-lockdown. The restrictions imposed limited movement outside, and all gyms were closed. These findings confirmed by previous studies (14,72,77). As we expected, the group with higher depression severity reported lower physical activity measured by the number of steps during pre and the lockdown, and vigorous activity during all periods with respect to the group with no or mild depression. Low physical activity is frequently observed in people with MDD (78,79).

### *Findings on participants with M*S

Depression is a common syndrome reported by people with MS (24,25), resulting in a low quality of life, increased fatigue and disability, and poor prognosis (25,80). The stressful situation due to COVID-19 restriction might have exacerbated depressive symptoms (23). However, in our study, participants with MS did not report depression changes during the three periods. As we observed for MDD, the group with severe depression maintained the level of depression during all three periods. Other studies confirmed this absence of changes in people from different countries with MS (81).

We observed changes in mean HR and HR variation during all day and night, resting periods during the day and night, and activity. The decrement was observed especially between pre-lockdown and lockdown, and between pre-lockdown and post-lockdown in daily mean HR and HR variation. Lower resting mean HR during the day may be related to a reduction of physical activity during the lockdown, we observed a diminution in the number of steps in the same period as we observed for the group with MDD.

The resting mean HR reduced during the day and night from pre-lockdown and during the lockdown, but the resting mean HR during the day increased between lockdown and post-lockdown. An elevated resting mean HR between the lockdown and post-lockdown may be caused by other factors such as psychological stress perception (38) acute respiratory infections (51,82) or an increased alcohol intake (83) et al. 2021). 21.8% of our participants with MS (total sample=399) reported major symptoms similar to COVID-19 symptoms (42) and found an association with HR parameters (51). Low HR variability was reported in people with MS (84), which may be caused by a significant increase in sympathetic cardiovascular tone (38) and can influence the course of the disease (39). The reduction of resting mean HR at night might be due to sleep alterations (38). A previous study also reported a sleep disruption in the same sample during the lockdown (14), which was confirmed in another populations (85). Future studies are needed to explore if HR parameters are altered due to other factors in this population.

Consistent with our hypothesis, individuals with MS reduced their activity as measured by the number of steps and minutes of light intensity of activity between pre and during lockdown, and increased between lockdown and post-lockdown. The group with lower depression reported a higher mean of steps during the pre-lockdown. No differences were found in the different intensities of activity across the periods. It is well known that people with MS suffer from fatigue and gait dysfunctions (86). This may be one of the reason why we observe changes only in light activity between pre and during lockdown but not for moderate and vigorous activity, and between pre and post-lockdown. The stressful situation caused by COVID-19 can add to the feeling of overwhelming fatigue (87). In the whole MS group, the online social activity remained stable, except for the social contacts. They may have also perceived feelings of loneliness (88). The lockdown increased social isolation for all people, but with different impacts on different groups.

### Strengths and limitations

The strengths of our study are that our sample was extensive in terms of observations and included participants from two different chronic diseases. Moreover, we had three periods: pre-lockdown, lockdown, and post-lockdown. Our main aim was to explore how the lockdown impacts two different health conditions by depression severity. Participants with MDD and MS answered a different number of depression questionnaires (PHQ-8). Nonetheless, we included the participants that completed a minimum of two questionnaires. The data extracted by the PPG are highly correlated with the parameters of HR variability removed by the ECG (89,90). However although wrist-worn devices seldom give access to the PPG signal but instead to HR series derived by proprietary algorithms. Participants came from four European countries; differences were explored in our previous work (14). We used an assessment based on real-time and conducted it in a natural environment. Our findings showed that the RADAR-base system could monitor psychosocial impact changes due to stressful life events. Various factors might have had an impact on the levels of depression: economic status, changes in contact with families, such as the restriction on seeing relatives face-to-face during the lockdown or the loss of physical demonstrations of affection, especially in countries that imposed stricter limitations than others, as was the case with Spain and Italy. Further decentralized research should consider these factors. Furthermore, as long as the technical details of Fitbit devices are not open to researchers, investigating the validity is difficult as it is not possible to know if the observed differences are results of the device itself or, more likely, differences in algorithms.

### Future directions

The RADAR-base tool allowed us to explore the risk factors for depression across diagnoses under the difficult circumstances caused by the pandemic national restrictions, future uncertainty and fear of the contagion, fear of one’s own death or that of their family members/friends. This situation could have aggravated the level of depression in people who already suffered from depression, increased the feeling of loneliness; and reduced physical activity, especially in people with moderate or severe depression. Low HR variation and increased resting mean HR appears to be a common pathophysiological factor across diagnosis. Future studies are required to explore how people with different depression severity react in stressful situations.

## Data Availability

All data produced in the present work are contained in the manuscript

## Contributors

SS (Sara Siddi), IG, RB, EP, KS, EG, JMH contributed to the study design. SS (Sara Siddi) and IG contributed to the data analysis, figures drawing, and manuscript writing.

MF, SS (Sara Siddi),GL, LF, PBWJH, OC, WK contributed to the data collection for the MDD group. CG, LL, MX, AZ AB, MM, PPS, MX, AZ AB, DCG, and BM contributed to the data collection for MS group. GC and MH led the MDD and MS group, respectively and contributed from the analysis plan for the present manuscript.

MH and VB: PIs for RADAR-CNS, funding, study design and oversight of data collection. FA, DR, DJ, YR, ZR, CN, VAN, PA contributed to the administrative, technical, and clinical support of the study. All authors contributed to the critical revision of the manuscript.

## Acknowledgment

Participants in the CIBER site came from following four clinical communities in Spain: Parc Sanitari Sant Joan de Déu Network services, Institut Català de la Salut, Institut Pere Mata, and Hospital Clínico San Carlos. Participant recruitment in Amsterdam was partially accomplished through Hersenonderzoek.nl, a Dutch online registry that facilitates participant recruitment for neuroscience studies Ref Hersenonderzoek.nl is funded by ZonMw-Memorabel project no 73305095003, a project in the context of the Dutch Deltaplan Dementie, Gieskes-Strijbis Foundation, the Alzheimer’s Society in the Netherlands and Brain Foundation Netherlands.

This paper represents independent research part funded by the National Institute for Health Research NIHR Maudsley Biomedical Research Centre at South London and Maudsley NHS Foundation Trust and King’s College London. The views expressed are those of the authors and not necessarily those of the NHS, the NIHR or the Department of Health and Social Care. We thank all the members of the RADAR-CNS patient advisory board for their contribution to the device selection procedures, and their invaluable advice throughout the study protocol design. This research was reviewed by a team with experience of mental health problems and their careers who have been specially trained to advise on research proposals and documentation through the Feasibility and Acceptability Support Team for Researchers FAST-R: a free, confidential service in England provided by the National Institute for Health Research Maudsley Biomedical Research Centre via King’s College London and South London and Maudsley NHS Foundation Trust. We thank all GLAD Study volunteers for their participation, and gratefully acknowledge the NIHR BioResource, NIHR BioResource centres, NHS Trusts and staff for their contribution. We also acknowledge NIHR BRC, King’s College London, South London and Maudsley NHS Trust and King’s Health Partners. We thank the National Institute for Health Research, NHS Blood and Transplant, and Health Data Research UK as part of the Digital Innovation Hub Programme. RADAR-MDD will be conducted per the Declaration of Helsinki and Good Clinical Practice, adhering to principles outlined in the NHS Research Governance Framework for Health and Social Care 2nd edition. Ethical approval has been obtained in London from the Camberwell St Giles Research Ethics Committee REC reference: 17/LO/1154, in London from the CEIC Fundacio Sant Joan de Deu CI: PIC-128-17 and in the Netherlands from the Medische Ethische Toetsingscommissie VUms METc VUmc registratienummer: 2018.012 – NL63557.029.17.

## Abbreviations

COVID-19: COronaVIrus Disease 19
HR: Heart rate
MDD: Major depressive disorder
MS: Multiple Sclerosis
mHR: mean HR
PPG: photoplethysmography
PHQ-8: 8-item Patient Health Questionnaire
RADAR-CNS: Remote assessment of disease and relapse - central nervous system
RADAR-MDD: Remote assessment of disease and relapse-major depressive disorder
RMT: Remote measurement technology
SARS-CoV-2: severe acute respiratory syndrome coronavirus 2
stdHR: Standard deviation Heart Rate
WHO: World Health Organization

## Supplementary materials

**Supplement S1:** Level of completeness of the data for the cohort of MDD and MS.

|  | MDD (N=255) |  | MS (N=213) |  |
| --- | --- | --- | --- | --- |
|  | Valid<br>N (%) | Observations<br>N | Valid<br>N (%) | Observations<br>N |
| **PHQ-8** | 255 (100) | 691 | 213 (100) | 479 |
| **HR variables** | 234 (91.76) | 571 | 193 (90.61) | 419 |
| **Steps** | 218 (85.49) | 561 | 192 ( 90.14 ) | 446 |
| **Sedentary, activity**<br>**light, moderate, and**<br>**vigorous** | 225 (88.23) | 593 | 199 ( 93.43 ) | 446 |
| **Social contacts** | 216 (84.70 ) | 512 | 157 ( 73.71) | 288 |
| **Social interactions** | 212 (83.14) | 491 | 172 (80.75 ) | 337 |

**Note:** Number of participants for each group who have data for the studied periods: pre-lockdown, during lockdown and post-lockdown. All variables of HR have the same N of Ids, as well activity (sedentary, light, moderate and vigorous intensity).

**Table S2:**
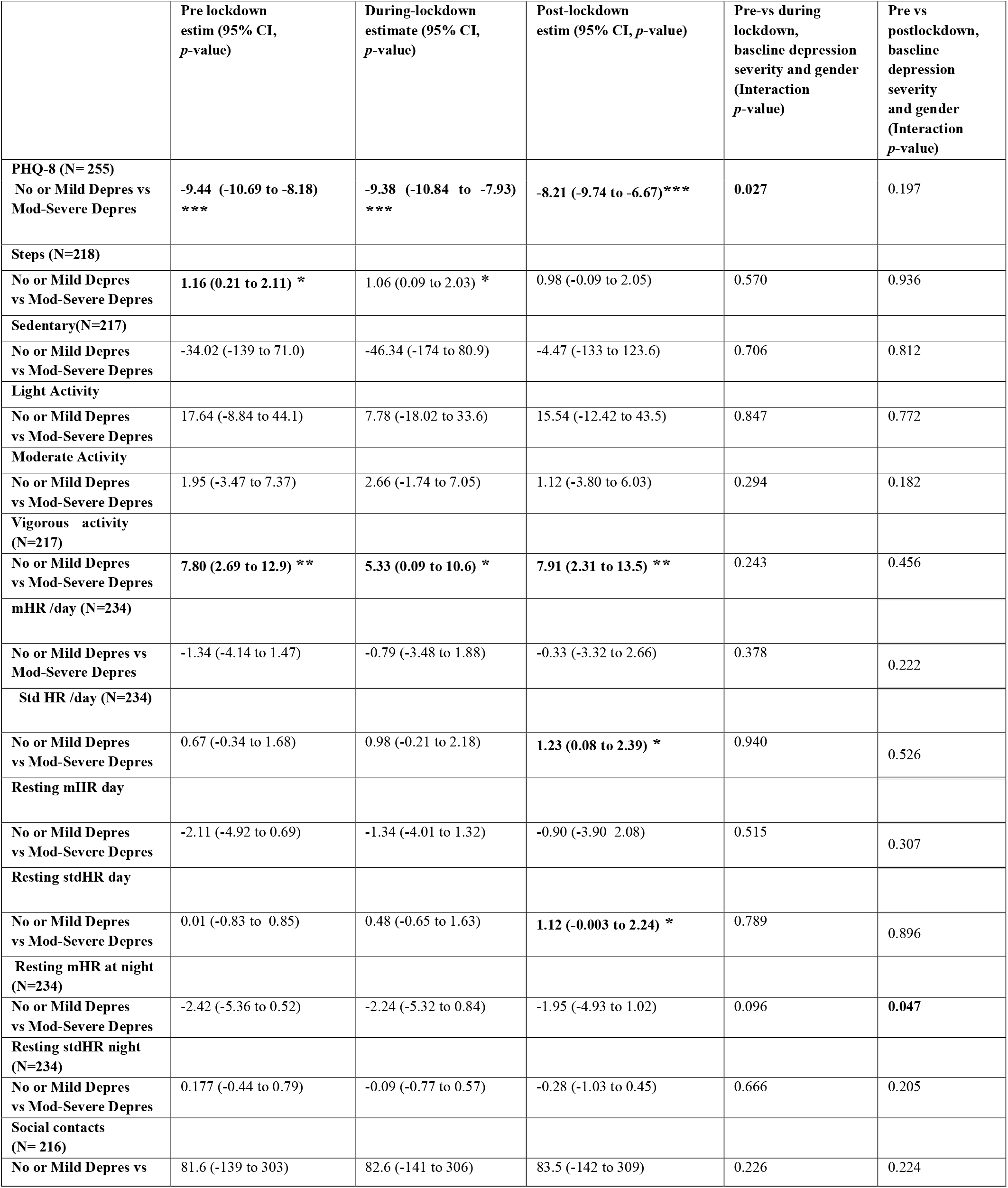
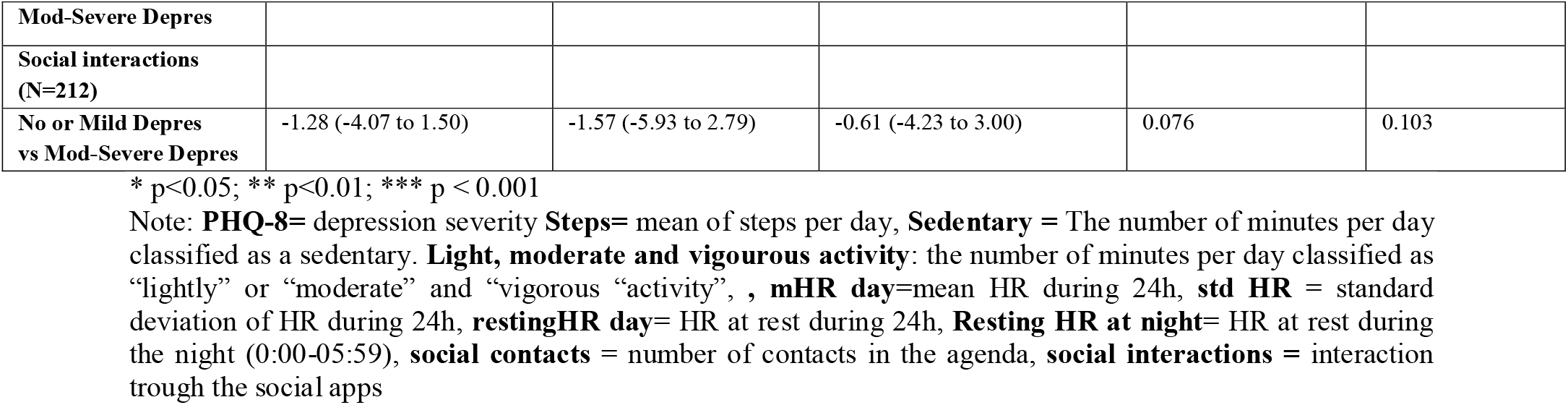
Estimated mean differences in each outcome between no or mild depression vs moderate or severe depression at each period and interaction with gender in MDD group.

|  | Pre lockdown<br>estim (95% CI,<br>*p*-value) | During-lockdown<br>estimate (95% CI,<br>*p*-value) | Post-lockdown<br>estim (95% CI, *p*-value) | Pre-vs during<br>lockdown,<br>baseline depression<br>severity and gender<br>(Interaction<br>*p*-value) | Pre vs<br>postlockdown,<br>baseline<br>depression<br>severity<br>and gender<br>(Interaction<br>*p*-value) |
| --- | --- | --- | --- | --- | --- |
| **PHQ-8 (N= 255)** |  |  |  |  |  |
| No or Mild Depres vs Mod-Severe Depres | **-9.44 (-10.69 to -8.18) \*\*\*** | **-9.38 (-10.84 to -7.93) \*\*\*** | **-8.21 (-9.74 to -6.67)\*\*\*** | **0.027** | 0.197 |
| **Steps (N=218)** |  |  |  |  |  |
| No or Mild Depres vs Mod-Severe Depres | **1.16 (0.21 to 2.11) \*** | 1.06 (0.09 to 2.03) * | 0.98 (-0.09 to 2.05) | 0.570 | 0.936 |
| **Sedentary(N=217)** |  |  |  |  |  |
| No or Mild Depres vs Mod-Severe Depres | -34.02 (-139 to 71.0) | -46.34 (-174 to 80.9) | -4.47 (-133 to 123.6) | 0.706 | 0.812 |
| **Light Activity** |  |  |  |  |  |
| No or Mild Depres vs Mod-Severe Depres | 17.64 (-8.84 to 44.1) | 7.78 (-18.02 to 33.6) | 15.54 (-12.42 to 43.5) | 0.847 | 0.772 |
| **Moderate Activity** |  |  |  |  |  |
| No or Mild Depres vs Mod-Severe Depres | 1.95 (-3.47 to 7.37) | 2.66 (-1.74 to 7.05) | 1.12 (-3.80 to 6.03) | 0.294 | 0.182 |
| **Vigorous activity (N=217)** |  |  |  |  |  |
| No or Mild Depres vs Mod-Severe Depres | **7.80 (2.69 to 12.9) \*\*** | **5.33 (0.09 to 10.6) \*** | **7.91 (2.31 to 13.5) \*\*** | 0.243 | 0.456 |
| **mHR /day (N=234)** |  |  |  |  |  |
| No or Mild Depres vs Mod-Severe Depres | -1.34 (-4.14 to 1.47) | -0.79 (-3.48 to 1.88) | -0.33 (-3.32 to 2.66) | 0.378 | 0.222 |
| **Std HR /day (N=234)** |  |  |  |  |  |
| No or Mild Depres vs Mod-Severe Depres | 0.67 (-0.34 to 1.68) | 0.98 (-0.21 to 2.18) | **1.23 (0.08 to 2.39) \*** | 0.940 | 0.526 |
| **Resting mHR day** |  |  |  |  |  |
| No or Mild Depres vs Mod-Severe Depres | -2.11 (-4.92 to 0.69) | -1.34 (-4.01 to 1.32) | -0.90 (-3.90 2.08) | 0.515 | 0.307 |
| **Resting stdHR day** |  |  |  |  |  |
| No or Mild Depres vs Mod-Severe Depres | 0.01 (-0.83 to 0.85) | 0.48 (-0.65 to 1.63) | **1.12 (-0.003 to 2.24) \*** | 0.789 | 0.896 |
| **Resting mHR at night (N=234)** |  |  |  |  |  |
| No or Mild Depres vs Mod-Severe Depres | -2.42 (-5.36 to 0.52) | -2.24 (-5.32 to 0.84) | -1.95 (-4.93 to 1.02) | 0.096 | **0.047** |
| **Resting stdHR night (N=234)** |  |  |  |  |  |
| No or Mild Depres vs Mod-Severe Depres | 0.177 (-0.44 to 0.79) | -0.09 (-0.77 to 0.57) | -0.28 (-1.03 to 0.45) | 0.666 | 0.205 |
| **Social contacts (N= 216)** |  |  |  |  |  |
| No or Mild Depres vs | 81.6 (-139 to 303) | 82.6 (-141 to 306) | 83.5 (-142 to 309) | 0.226 | 0.224 |
| **Mod-Severe Depres** |  |  |  |  |  |
| **Social interactions**<br>(N=212) |  |  |  |  |  |
| **No or Mild Depres**<br>**vs Mod-Severe Depres** | -1.28 (-4.07 to 1.50) | -1.57 (-5.93 to 2.79) | -0.61 (-4.23 to 3.00) | 0.076 | 0.103 |

\* p<0.05; \*\* p<0.01; \*\*\* p < 0.001
Note: **PHQ-8**= depression severity **Steps**= mean of steps per day, **Sedentary** = The number of minutes per day classified as a sedentary. **Light, moderate and vigorous activity**: the number of minutes per day classified as “lightly” or “moderate” and “vigorous “activity”, , **mHR day**=mean HR during 24h, **std HR** = standard deviation of HR during 24h, **restingHR day**= HR at rest during 24h, **Resting HR at night**= HR at rest during the night (0:00-05:59), **social contacts** = number of contacts in the agenda, **social interactions** = interaction trough the social apps

**Table S3:**
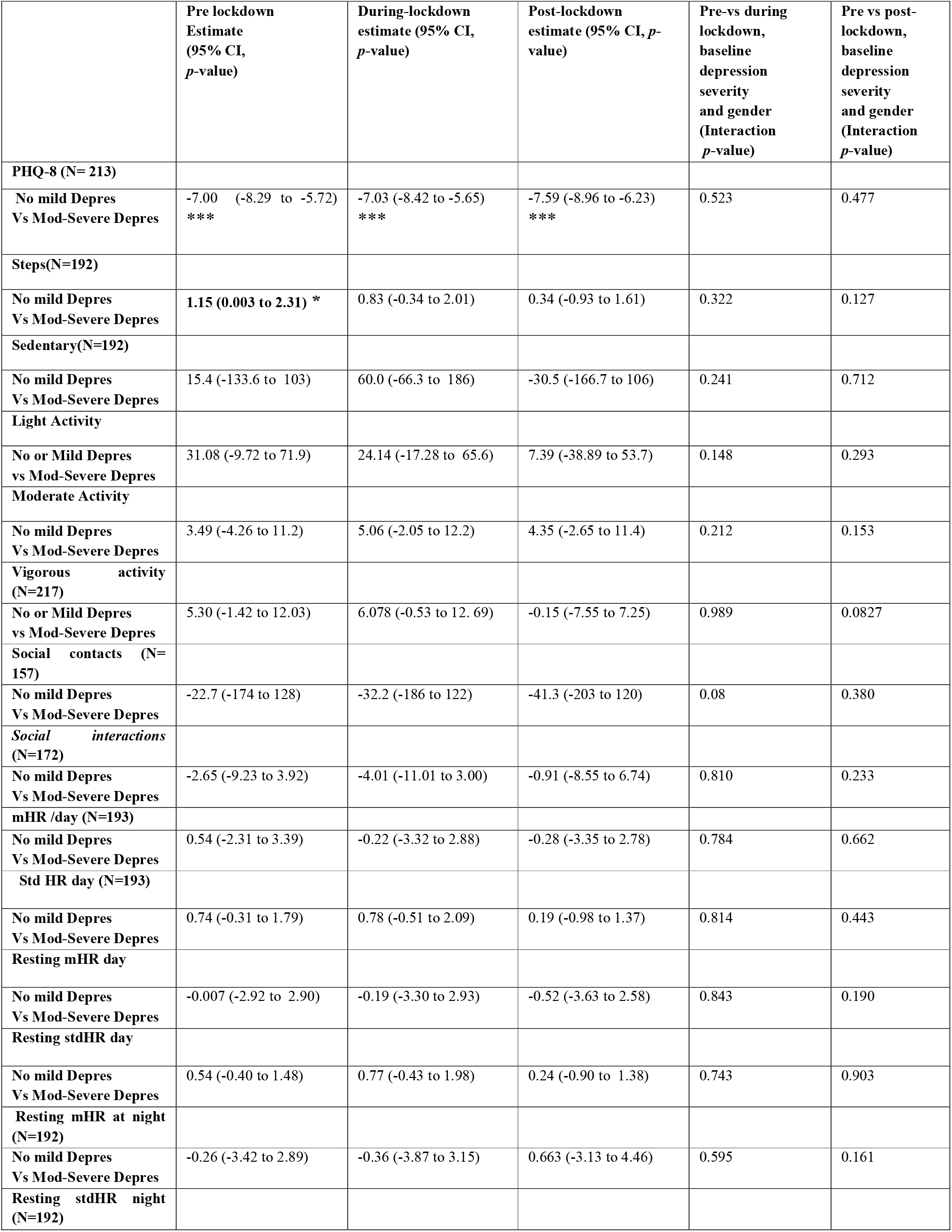
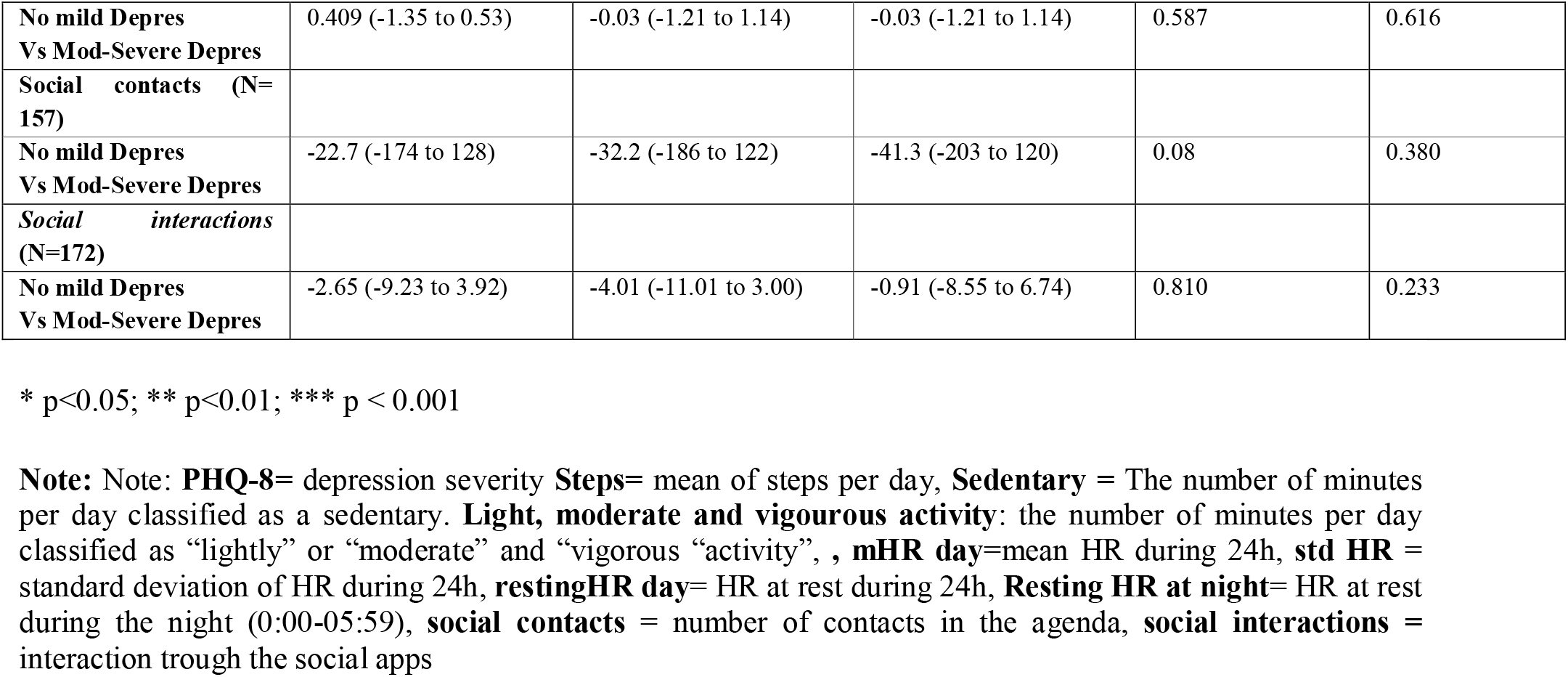
Estimated mean differences in each outcome between no or mild depression (vs moderate or severe depression at each period and interaction with gender in MS group.

